# Nationwide incidence of sarcomas and connective tissue tumors of intermediate malignancy over four years using an expert pathology review network

**DOI:** 10.1101/2020.06.19.20135467

**Authors:** Gonzague de Pinieux, Marie Karanian-Philippe, Francois Le Loarer, Sophie Le Guellec, Sylvie Chabaud, Philippe Terrier, Corinne Bouvier, Maxime Battistella, Agnès Neuville, Yves-Marie Robin, Jean-Francois Emile, Anne Moreau, Frederique Larousserie, Agnes Leroux, Nathalie Stock, Marick Lae, Francoise Collin, Nicolas Weinbreck, Sebastien Aubert, Florence Mishellany, Céline Charon-Barra, Sabrina Croce, Laurent Doucet, Isabelle Quintin-Rouet, Marie-Christine Chateau, Celine Bazille, Isabelle Valo, Bruno Chetaille, Nicolas Ortonne, Anne Gomez-Brouchet, Philippe Rochaix, Anne De Muret, Jean-Pierre Ghnassia, Lenaig Mescam-Mancini, Nicolas Macagno, Isabelle Birtwisle-Peyrottes, Christophe Delfour, Emilie Angot, Isabelle Pommepuy, Dominique Ranchere-Vince, Claire Chemin-Airiau, Myriam Jean-Denis, Yohan Fayet, Jean-Baptiste Courrèges, Nouria Mesli, Juliane Berchoud, Maud Toulmonde, Antoine Italiano, Axel Le Cesne, Nicolas Penel, Francoise Ducimetiere, Francois Gouin, Jean-Michel Coindre, Jean-Yves Blay, on behalf of the NETSARC/REPPS/RESOS and French Sarcoma Group-Groupe d’Etude des Tumeurs Osseuses (GSF-GETO) networks (Supplementary data document 1)

**Author notes:** **Correspondence to** Prof J.-Y Blay, Department of Medical Oncology, Centre Léon Bérard, 28 rue Laënnec, 69373 Lyon Cedex 08, & Université Claude Bernard Lyon I France. the first 3 authors (alphabetical order) contributed equally to this work. the last 3 authors contributed equally to this work.

## Abstract

**Background:** Since 2010, NETSARC and RREPS collected and reviewed prospectively all cases of sarcomas and tumors of intermediate malignancy (TIM) nationwide.

**Methods:** The nationwide incidence of sarcoma or TIM (2013-2016), confirmed by expert pathologists using WHO classification are presented. Yearly variations and correlation with published clinical trials was analyzed.

**Results:** 139 histological subtypes are reported among the 25172 patients with sarcomas (n=18710, 64%) or TIM (n=6460, 36%), respectively n=5838, n=6153, n=6654, and n=6527 yearly from 2013 to 2016. Over these 4 years, the yearly incidence of sarcomas and TIM was therefore 79.7, 24.9 and 95.1/10^6^/year, above that previously reported. GIST, liposarcoma, leiomyosarcomas, undifferentiated sarcomas represented 13%, 13%, 11% and 11% of tumors. Only GIST, as a single entity had a yearly incidence above 10/million/year. There were respectively 30, 63 and 66 different histological subtypes of sarcomas or TIM with an incidence ranging from 10 to 1/10^6^, 1-0.1/10^6^, or < 0.1/10^6^/year respectively. The 2 later “incidence groups” included 21% of the patients. The incidence of 8 histotypes varied significantly over this 4 years. Patients with tumors with an incidence above 1/10^6^per year have significantly higher numbers of dedicated published phase III and phase II clinical trials (p<10^−6^).

**Conclusions:** This nationwide registry of sarcoma patients with histology reviewed by sarcoma experts shows that the incidence of sarcoma and TIM is higher than reported, and that tumors with an incidence<10^6^/year have a much lower access to clinical trials.

## Introduction

Sarcomas is an heterogeneous group of rare connective tissue cancers, with variable clinical presentations, and a reported incidence thought to be close to 2/10^5^/year 15 years ago, which has more recently been reported to range from 3 to 7 /10^5^/year (1-16).

The reported incidence of sarcoma varies considerably across countries and according to the date of analysis (2-16). The overall incidence of sarcoma is therefore not precisely known, and even less so that of individual histological subtypes, which are not unfrequently misdiagnosed. Indeed, because of their rarity, sarcoma are initially misclassified in up to 30% of cases (1,5,6,11,17). As a consequence patients with sarcomas may not treated according to clinical practice guidelines (1-21). In all clinical practice guidelines, it is recommended that the diagnosis of sarcoma should be confirmed by an expert pathologist. In general, management of sarcoma patients should be performed by a dedicated multidisciplinary team, including expert pathologists and surgeons, treating a large number of patients (5-7). It was recently reported that central pathology review of sarcoma cases is cost effective, reducing both morbidity, mortality and cost of management (22,23).

Since 2010, the French National Cancer Institute (INCa) funded pathology and clinical networks for sarcoma called RREPS and NETSARC, subsequently joined by RESOS focused on bone sarcomas, to improve the quality of management of sarcoma patients. Initially, the network of 23 expert reference centers for pathology (RRePS) was in charge of the mandatory histological review for each suspected case of sarcoma nationwide. These networks have merged since 2019 in a single NETSARC+ network. The common database (netsarc.org) gathering all cases of sarcoma presented to MDTB was created and implemented, collected data on the diagnostic, therapeutic management, and the clinical outcome in terms of relapse and survival. From Jan 1^st^ 2010, this database prospectively included over 59000 patients with sarcoma or tumor of intermediate malignancy. Since 2013, the overall accrual in the database reached a plateau, suggesting that the closest to exhaustive collection of cases in this country was obtained.

The incidence of sarcoma and TIM has seldom been reported in exhaustive nationwide series with organized reference center pathology review. We report here on the incidence of the different histological subtypes of sarcomas and TIM reported in the NETSARC+ database from 2013 to 2016.

## Patient, material and methods

### The NETSARC+ network and the referral of the pathology sample to the network of experts

The RREPS (pathology, NETSARC (clinical management), and RESOS (bone sarcoma) networks which gathered experts from the same centers were merged into the NETSARC+ network in 2019. The organization of these networks has been reported previously (24,25): each RREPS and NETSARC center organizes a multidisciplinary tumor board (MDTB) gathering sarcoma specialized pathologist(s), radiologist(s), surgeon(s), radiation oncologist(s), medical oncologist(s), and often molecular biologist(s), orthopedist(s), pediatrician(s).

Since 2010, it is mandatory for the primary pathologist to refer all suspected cases of sarcomas or TIM to one of the reference centers. In addition, if this has not been done previously, pathology review is will be requested for any clinical case referred to one of the 26 multidisciplinary tumor board of NETSARC without initial prior central pathology review, thus ensuring a double check. These two modes of entry improve the rate of review of the sarcoma/TIM samples nationwide. All sarcoma/TIM or suspected sarcoma/TIM patient cases presented to the MDTB of all 26 centers were recorded in the electronic online database, by a dedicated team of Clinical research assistant (CRAs), supervised by the Coordinating centers (Centre Leon Bérard, Gustave Roussy, Institut Bergonié, CHU Tours, CHU Nantes). Patient files may be presented before any diagnostic procedure, before initial biopsy, before primary surgery, after primary surgery, at relapse, and/or in case of a possible inclusion in a clinical trial as previously described (24,25). Patients and treatment data were prospectively included and regularly updated by the dedicated study coordinators.

### The RREPS/NETSARC Database

The RREPS/NETSARC database may therefore enable to describe as exhaustively as possible the incident and prevalent population of sarcoma patients in France. Of note, the database includes a limited set of data, on purpose, describing patients and tumor characteristics, surgery, relapse and survival (24, 25), centers performing the first resection, as well as potential secondary surgery types and sites, the final quality of resection, etc…

Of note, about 24% of patients in the database discussed in a NETSARC MDTB had a diagnosis which was not that of a sarcoma/TIM (e.G. lipoma, carcinoma, lymphoma…). Again, it is important to note that in NETSARC, patients with suspected sarcoma/TIM can enter the process of MDTB either through the pathology network, or directly by the physician, leading both to a final MDTB review after central pathology confirmation.

All data presented here were extracted from the NETSARC.org database accessible online for a period of 4 years between 2013 and 2016. These 4 years were selected since: 1) the yearly incidence of sarcoma and TIM started to plateau since 2013, and 2) data monitoring and implementation is still ongoing for year 2017 and later.

### Presentation of the data

The WHO classification is used to describe the histological subtypes in the database (1). The number of patients for each individual histological subtype of sarcoma or TIM per year, from 2013 to 2016, is therefore presented in these tables. To facilitate the comparison with other databases using previous classifications, the incidence for groups of tumors are also presented in the Tables, when they are clinically relevant (e.g. uterine sarcoma), or used as entities for clinical trials (e.g. liposarcomas, leiomyosarcoma, solitary fibrous tumors, giant cell tumors of the bone…). To estimate the incidence of these tumors, we used the official number of French citizens in the years 2013 to 2016, which were respectively 65.56, 66.13, 66.42 and 66.60 millions inhabitants.

### Matching histotypes with published clinical trials

Each individual histotype was screened to identify a dedicated clinical trial within the Pubmed database. The name of the entity (e.g. angiosarcoma, pleomorphic liposarcoma…) was used in the interrogation, together with a filter on clinical trial, adding ≪ phase III ≫, ≪ randomized phase II ≫, or ≪ phase II ≫. Pubmed was interrogated between Jan 15 and Jan 30 2020.

### Statistical analyses

The number of patient per year with the different histotypes is presented in tables. To analyze the variation of incidence over the 4 years, an ANOVA procedure was used for the whole dataset. Histotypes with a significant variation in the period of observation are detailed. The comparison of the frequency of published clinical trials per histological subtypes or groups of subtypes was performed using the chi square or Fisher’s exact test. All statistical tests were two-sided. All statistical analyses were performed using SPSS (v 23.0) (IBM, Paris France).

### Funding

NetSARC (INCA & DGOS) and RREPS (INCA & DGOS), RESOS (INCA & DGOS) and LYRICAN (INCA-DGOS-INSERM 12563), Institut Convergence PLASCAN (17-CONV-0002), Association DAM’s, Ensemble contre Le GIST, Eurosarc (FP7-278742), la Fondation ARC, Infosarcome, InterSARC (INCA), LabEx DEvweCAN (ANR-10-LABX-0061), Ligue de L’Ain contre le Cancer, La Ligue contre le Cancer, EURACAN (EC 739521) funded the study. The funders had no role in the study design, data collection, data analysis, data interpretation, or writing of the report.

## Results

### Incidence of sarcoma and TIM in NETSARC

Table 1 to 3 present the incidence of the individual histological subtypes of soft tissue, visceral, bone sarcomas or connective tissue tumors included in the NETSARC+ databases from 2013 to 2016, the first 4 years for which it was considered close to exhaustive.

**Table 1:** incidence of soft tissue and visceral sarcomas

|  | 2013 | 2014 | 2015 | 2016 | Total | Incidence<br>/10<sup>6</sup>/year |
| --- | --- | --- | --- | --- | --- | --- |
| **Adipocytic tumours** | **744** | **821** | **817** | **865** | **3247** | **12,299** |
| Atypical lipomatous tumour /<br>well-differentiated liposarcoma | 289 | 304 | 314 | 357 | 1266 | 4,795 |
| Liposarcoma – dedifferentiated | 304 | 344 | 341 | 356 | 1345 | 5,095 |
| **Myxoid Round Cell LPS** | **99** | **106** | **108** | **96** | **409** | **1,549** |
| Liposarcoma - myxoid | 81 | 90 | 95 | 89 | 355 | 1,345 |
| Liposarcoma - round cell | 18 | 16 | 13 | 7 | 54 | 0,205 |
| Liposarcoma - pleomorphic | 31 | 41 | 36 | 31 | 139 | 0,527 |
| Lipomatous spindle cell/pleomorphic | 0 | 1 | 0 | 0 | 1 | 0,004 |
| Liposarcoma NOS | 21 | 25 | 17 | 22 | 85 | 0,322 |
| Liposarcoma - mixed type | 0 | 0 | 1 | 1 | 2 | 0,008 |
| **Fibroblastic & myofibroblastic tumours** | **1041** | **1047** | **1115** | **1147** | **4349** | **16,473** |
| Desmoid fibromatosis | 307 | 295 | 357 | 381 | 1340 | 5,072 |
| Lipofibromatosis | 3 | 0 | 5 | 0 | 8 | 0,030 |
| Giant cell Fibroblastoma | 2 | 4 | 4 | 1 | 11 | 0,042 |
| Dermatofibrosarcoma Protuberans | 261 | 270 | 258 | 251 | 1040 | 3,939 |
| **Solitary fibrous tumour (all)** | **210** | **222** | **242** | **252** | **925** | **3,504** |
| Solitary fibrous tumor | 166 | 178 | 193 | 214 | 751 | 2,845 |
| High risk SFT | 44 | 43 | 49 | 38 | 174 | 0,659 |
| Inflammatory myofibroblastic Tum. | 32 | 39 | 33 | 41 | 145 | 0,549 |
| Low grade Myofibroblastic Sarc. | 3 | 5 | 3 | 2 | 13 | 0,049 |
| Myxoinflammatory Fibroblastic Sarc. | 6 | 6 | 6 | 5 | 23 | 0,087 |
| Infantile fibrosarcoma | 3 | 2 | 1 | 4 | 10 | 0,038 |
| Adult fibrosarcoma | 11 | 4 | 9 | 4 | 28 | 0,106 |
| Myxofibrosarcoma | 162 | 160 | 152 | 156 | 630 | 2,386 |
| Low grade fibromyxoid sarcoma | 33 | 30 | 35 | 38 | 136 | 0,515 |
| Sclerosing epithelioid fibrosarcoma | 8 | 11 | 10 | 12 | 41 | 0,155 |
| **So-called fibrohistiocytic tumours** | **29** | **16** | **37** | **24** | **106** | **0,402** |
| Intermediate fibrohistiocytic tumors | 0 | 0 | 2 | 3 | 5 | 0,019 |
| Malignant tenosynovial giant cell tum. | 1 | 0 | 0 | 1 | 2 | 0,008 |
| Plexiform fibrohistiocytic tumors | 7 | 7 | 9 | 6 | 29 | 0,110 |
| Giant cell tumour of soft tissue | 21 | 9 | 26 | 14 | 70 | 0,265 |
| **Vascular tumours** | **398** | **377** | **381** | **364** | **1520** | **5,758** |
| Retiform hemangio-endothelioma | 1 | 3 | 3 | 2 | 9 | 0,034 |
| Papillary intralymphatic<br>angioendothelioma | 0 | 0 | 0 | 1 | 1 | 0,004 |
| Composite hemangioendothelioma | 1 | 1 | 1 | 0 | 3 | 0,011 |
| Kaposi sarcoma | 191 | 165 | 162 | 145 | 663 | 2,511 |
| Kaposiform hemangioendothelioma | 1 | 1 | 1 | 1 | 4 | 0,015 |
| Pseudomyogenic hemangioendothelioma | 1 | 3 | 0 | 2 | 6 | 0,023 |
| Epithelioid hemangioEndothelioma | 27 | 20 | 30 | 23 | 100 | 0,379 |
| Angiosarcoma | 176 | 183 | 182 | 187 | 728 | 2,758 |
| Intermediate vascular tumours | 0 | 1 | 2 | 3 | 6 | 0,023 |
| **Pericytic (perivascular) tumours** | **4** | **4** | **1** | **1** | **10** | **0,038** |
| Malignant glomus tumour |  |  |  |  |  |  |
| **Smooth muscle (SM) tumours** | **646** | **698** | **669** | **666** | **2679** | **10,148** |
| SM tumor of undetermined malignancy | 20 | 47 | 23 | 32 | 122 | 0,462 |
| Metastatic leiomyoma | 0 | 0 | 0 | 2 | 2 | 0,008 |
| Leiomyosarcoma | 247 | 263 | 287 | 297 | 1094 | 4,144 |
| Leiomyosarcoma -differentiated | 245 | 243 | 240 | 217 | 945 | 3,580 |
| Leiomyosarcoma – poorly differentiated | 134 | 145 | 119 | 118 | 516 | 1,955 |
| **Skeletal muscle sarcoma (RMS)** | **145** | **157** | **173** | **133** | **608** | **2,303** |
| **Embryonal RMS** | **50** | **45** | **60** | **34** | **179** | **0,678** |
| Embryonal RMS sarcoma - botryoid type | 8 | 6 | 6 | 3 | 23 | 0,087 |
| Embryonal rhabdomyosarcoma usual type | 35 | 31 | 47 | 24 | 137 | 0,519 |
| Embryonal rhabdomyosarcoma spindle cell | 7 | 8 | 7 | 7 | 29 | 0,110 |
| Alveolar RMS | 27 | 36 | 35 | 25 | 123 | 0,466 |
| Pleomorphic RMS | 28 | 38 | 42 | 36 | 144 | 0,545 |
| Sclerosing RMS | 2 | 3 | 3 | 3 | 11 | 0,042 |
| Spindle cell RMS | 13 | 8 | 9 | 9 | 39 | 0,148 |
| Adult spindle cell RMS | 0 | 0 | 1 | 4 | 5 | 0,019 |
| RMS NOS | 21 | 25 | 23 | 19 | 88 | 0,333 |
| Ectomesenchymoma: Mal. mesenchymoma | 4 | 2 | 0 | 3 | 9 | 0,034 |
| **Gastrointestinal stromal tumors (GIST).** | **736** | **792** | **913** | **831** | **3272** | **12,394** |
| **Chondro-osseous tumours** |  |  |  |  |  |  |
| Extraskeletal osteosarcoma | **25** | **25** | **32** | **14** | **96** | **0,364** |
| **Peripheral nerve sheath tumours** | **75** | **68** | **69** | **74** | **286** | **1,083** |
| MPNST - epithelioid type | 0 | 2 | 1 | 3 | 6 | 0,023 |
| MPNST - usual type | 36 | 7 | 14 | 28 | 85 | 0,322 |
| Malignant peripheral nerve sheath tumour | 36 | 55 | 47 | 35 | 173 | 0,655 |
| Malignant Triton tumour | 0 | 2 | 3 | 5 | 10 | 0,038 |
| Malignant granular cell Tumour | 3 | 2 | 4 | 0 | 9 | 0,034 |
| Malignant perineurioma | 0 | 0 | 0 | 3 | 3 | 0,011 |

**Table 1 (part 2): incidence of soft tissue and visceral sarcomas**
|  | 2013 | 2014 | 2015 | 2016 | Total | Incidence<br>/10e6/year |
| --- | --- | --- | --- | --- | --- | --- |
| **Tumours of uncertain differentiation** |  |  |  |  |  |  |
| Atypical fibroxanthoma | 114 | 107 | 89 | 119 | 429 | 1,625 |
| Angiomatoid fibrous histiocytoma | 9 | 15 | 10 | 9 | 43 | 0,163 |
| Ossifying fibromyxoid Tumour | 7 | 7 | 5 | 13 | 32 | 0,121 |
| **Myoepithelioma, myoepithelial carcinoma,**<br>**& mixed tumour** | **31** | **26** | **18** | **18** | **96** | **0,364** |
| Myoepithelioma | 30 | 26 | 15 | 14 | 85 | 0,322 |
| Malignant myoepithelial Tumour | 0 | 0 | 1 | 1 | 2 | 0,008 |
| Mixed tumour | 1 | 0 | 2 | 3 | 6 | 0,023 |
| Haemosiderotic fibrolipomatous tumour | 0 | 2 | 0 | 7 | 9 | 0,034 |
| Phosphaturic mesenchymal tumour | 0 | 1 | 2 | 2 | 5 | 0,019 |
| NTRK-rearranged spindle cell neoplasm (emerging) | Not reported in NETSARC (so far) |  |  |  |  |  |
| **Synovial sarcoma** | **103** | **101** | **133** | **105** | **442** | **1,674** |
| Synovial sarcoma – NOS | 23 | 18 | 29 | 21 | 91 | 0,345 |
| Synovial sarcoma - biphasic | 11 | 19 | 23 | 17 | 70 | 0,265 |
| Synovial sarcoma – monophasic | 60 | 57 | 67 | 60 | 244 | 0,924 |
| Synovial sarcoma - poorly Differentiated | 9 | 7 | 14 | 7 | 37 | 0,140 |
| **Epithelioid sarcoma (all)** | **29** | **30** | **28** | **33** | **120** | **0,455** |
| Epithelioid sarcoma | 23 | 28 | 25 | 22 | 98 | 0,371 |
| Undifferentiated epithelioid sarcoma | 6 | 2 | 3 | 11 | 22 | 0,083 |
| Alveolar soft part sarcoma | 10 | 7 | 8 | 6 | 31 | 0,117 |
| Clear cell sarcoma of soft tissue | 13 | 16 | 26 | 16 | 71 | 0,269 |
| Extraskeletal myxoid chondrosarcoma | 15 | 12 | 20 | 11 | 58 | 0,220 |
| Desmoplastic small round cell tumour | 14 | 9 | 12 | 17 | 52 | 0,197 |
| Extrarenal rhabdoid tumour | 6 | 13 | 16 | 16 | 51 | 0,193 |
| SMARCA4-deficient thoracic sarcoma | 0 | 0 | 6 | 9 | 15 | 0,057 |
| **PEComa, including angiomylipoma** | **13** | **27** | **15** | **29** | **86** | **0,326** |
| PECOMA - NOS | 13 | 25 | 11 | 18 | 67 | 0,254 |
| Malignant PECOMA | 0 | 2 | 4 | 13 | 19 | 0,072 |
| Intimal sarcoma | 14 | 12 | 11 | 9 | 46 | 0,174 |
| **Undifferentiated sarcoma (all)** | **566** | **627** | **784** | **740** | **2717** | **10,292** |
| Undifferentiated pleomorphic sarcoma | 290 | 367 | 470 | 429 | 1556 | 5,894 |
| Undifferentiated sarcoma | 110 | 87 | 154 | 79 | 430 | 1,629 |
| Undifferentiated sarcoma -NOS | 125 | 130 | 111 | 57 | 423 | 1,602 |
| Undifferentiated spindle cell sarcoma | 41 | 43 | 49 | 175 | 308 | 1,167 |
| Low grade sinonasal sarcoma | 2 | 0 | 0 | 3 | 5 | 0,019 |
| Melanotic neuroectodermal tumour infancy | 0 | 0 | 0 | 1 | 1 | 0,004 |
| **Phyllode sarcoma** | **32** | **25** | **46** | **35** | **138** | **0,523** |
| **Sarcomas or TIM NOS** |  |  |  |  |  |  |
| - Sarcoma NOS | 166 | 197 | 259 | 189 | 809 | 3,064 |
| - Tumors of intermediate malignancy | 7 | 15 | 13 | 17 | 52 | 0,197 |

From 2013 to 2016, a total of 25172 incident patients were included in the database (Table 1 and 2), with n=5838, n=6153, n=6654, and n=6527 new patients for each year.

**Table 2:**
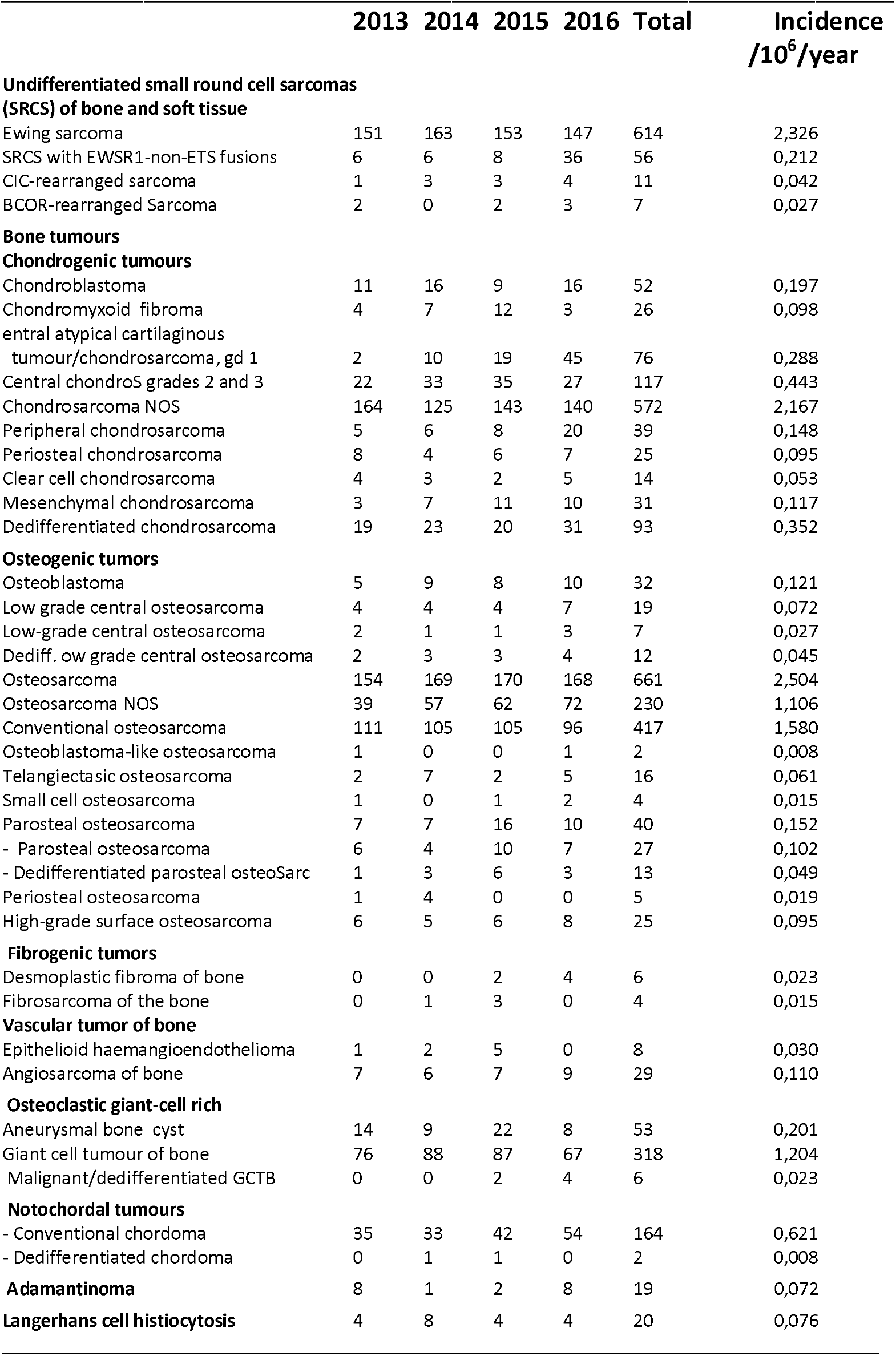
Incidence of bone sarcomas in NETSARC+ (2013-2016)

The NETSARC database includes 156 individual tumors or groups of sarcoma/TIM, 31 groups of sarcomas/TIM (e.g. « liposarcoma ») and 125 distinct individual histological subtypes of sarcomas or TIM (Table 1-3). Twelve additional histological subtypes of bone sarcomas (leiomyosarcomas, synovial etc) were also distinguished in this work (Table 3). Finally, Table 3 also presents the incidence of sarcomas diagnosed in patients with reported genetic predispositions.

**Table 3:** Incidence of uterine and rare bone sarcomas & genetic syndromes

|  | 2013 | 2014 | 2015 | 2016 | Total | Incidence<br>/10e6/year |
| --- | --- | --- | --- | --- | --- | --- |
| **Uterine sarcoma** | **242** | **311** | **285** | **300** | **1138** | **4,311** |
| **Endometrial stromal sarcoma, low grade** | **57** | **64** | **55** | **62** | **238** | **0,902** |
| Endometrial stromal nodule | 2 | 1 | 5 | 8 | 16 | 0,061 |
| Endometrial stromal sarcoma | 0 | 3 | 2 | 5 | 10 | 0,038 |
| Endometrial stromal sarcoma-low grade | 55 | 60 | 48 | 49 | 212 | 0,803 |
| Endometrial stromal sarcoma - high-grade | 1 | 5 | 13 | 22 | 41 | 0,155 |
| Adenosarcoma | 28 | 35 | 42 | 51 | 156 | 0,591 |
| Undifferentiated uterine sarcoma | 37 | 49 | 38 | 17 | 141 | 0,534 |
| Uterine tumour resembling ovarian sex cord | 5 | 1 | 4 | 7 | 17 | 0,064 |
| **Uterine leiomyosarcoma**<br>(extracted from the LMS group above) | **114** | **157** | **133** | **141** | **545** | **2,064** |
| **Rare bone sarcomas**<br>(extracted from the histological groups in Table 1) |  |  |  |  |  |  |
| **All undifferentiated sarcoma of bone** | **36** | **31** | **38** | **47** | **152** | **0,576** |
| Undifferentiated pleomorphic sarcoma of bone | 16 | 20 | 12 | 21 | 69 | 0,261 |
| Undifferentiated sarcoma | 16 | 11 | 21 | 8 | 56 | 0,212 |
| Undifferentiated spindle cell sarcoma | 4 | 0 | 5 | 17 | 26 | 0,098 |
| Undifferentiated epithelioid sarcoma | 0 | 0 | 0 | 1 | 1 | 0,004 |
| Leiomyosarcoma of bone | 11 | 15 | 5 | 9 | 40 | 0,152 |
| Synovial sarcoma of bone | 4 | 2 | 2 | 1 | 9 | 0,034 |
| Rhabdomyosarcoma of bone | 2 | 2 | 1 | 4 | 9 | 0,034 |
| BCOR Sarcoma of bone | 1 | 0 | 2 | 3 | 6 | 0,023 |
| Myoepithelioma of bone | 1 | 1 | 1 | 1 | 4 | 0,015 |
| Liposarcoma of bone | 0 | 2 | 0 | 2 | 4 | 0,015 |
| Other histological subtypes of bone sarcomas | 33 | 42 | 46 | 50 | 171 | 0,648 |
| **Genetic predisposition of soft tissue and bone or HIV** |  |  |  |  |  |  |
| Enchondromatosis | 5 | 2 | 6 | 4 | 17 | 0,064 |
| Li Fraumeni syndrome | 3 | 3 | 4 | 4 | 14 | 0,053 |
| Retinoblastoma | 1 | 0 | 3 | 1 | 5 | 0,019 |
| Multiple osteochondroma | 2 | 2 | 11 | 5 | 20 | 0,076 |
| Neurofibromatosis | 28 | 28 | 24 | 25 | 105 | 0,398 |
| Rothmund-Thomson | 0 | 1 | 0 | 0 | 1 | 0,004 |
| HIV | 4 | 12 | 10 | 6 | 32 | 0,121 |
| Other immunosuppression | 3 | 13 | 4 | 5 | 25 | 0,095 |

With an official number of the French population of 65.56, 66.13, 66.42 and 66.60 millions inhabitants in these 4 years, the estimated incidence of sarcomas and tumors of intermediate malignancy from 2013 to 2016 was 89.05, 93.04, 100.18, and 98.00 per million inhabitants respectively. Over these 4 years, the estimated yearly incidence of sarcomas and TIM was therefore 95,10/10^6^/year.

There were 18710 (64%) patients with sarcomas (incidence 70.87/10^e^/year) and 6460 (36%, 24.47/10^6^/year) patients with TIM.

The observed overall incidence of sarcoma and TIMs is therefore above that previously reported (1-15).

### Over 100-fold difference in incidence in different sarcoma histotypes

Figure 1 presents the individual histotypes and relevant groups of histotypes (eg liposarcoma, leiomyosarcoma, unterine sarcomas) ordered by incidence. GIST, liposarcoma, leiomyosarcomas, undifferentiated sarcomas represented 13%, 13%, 11% and 11% of all sarcomas (47% all 4 together). Only gastrointestinal stromal tumors, if considered as single entities, exceeded a yearly incidence above 10/10^6^/ year (Figure 1). The other histological types of sarcomas with a yearly incidence above 10/10^6^/year are histotypes groups 1) all liposarcomas, 2) all smooth muscle tumors, 3) all undifferentiated sarcomas, and 4) all fibroblastic or myofibroblastic tumors lumped together. This later group a group is not clinically homogenous and usually not considered as a specific entity in clinical trials or retrospective studies.

**Figure 1:**
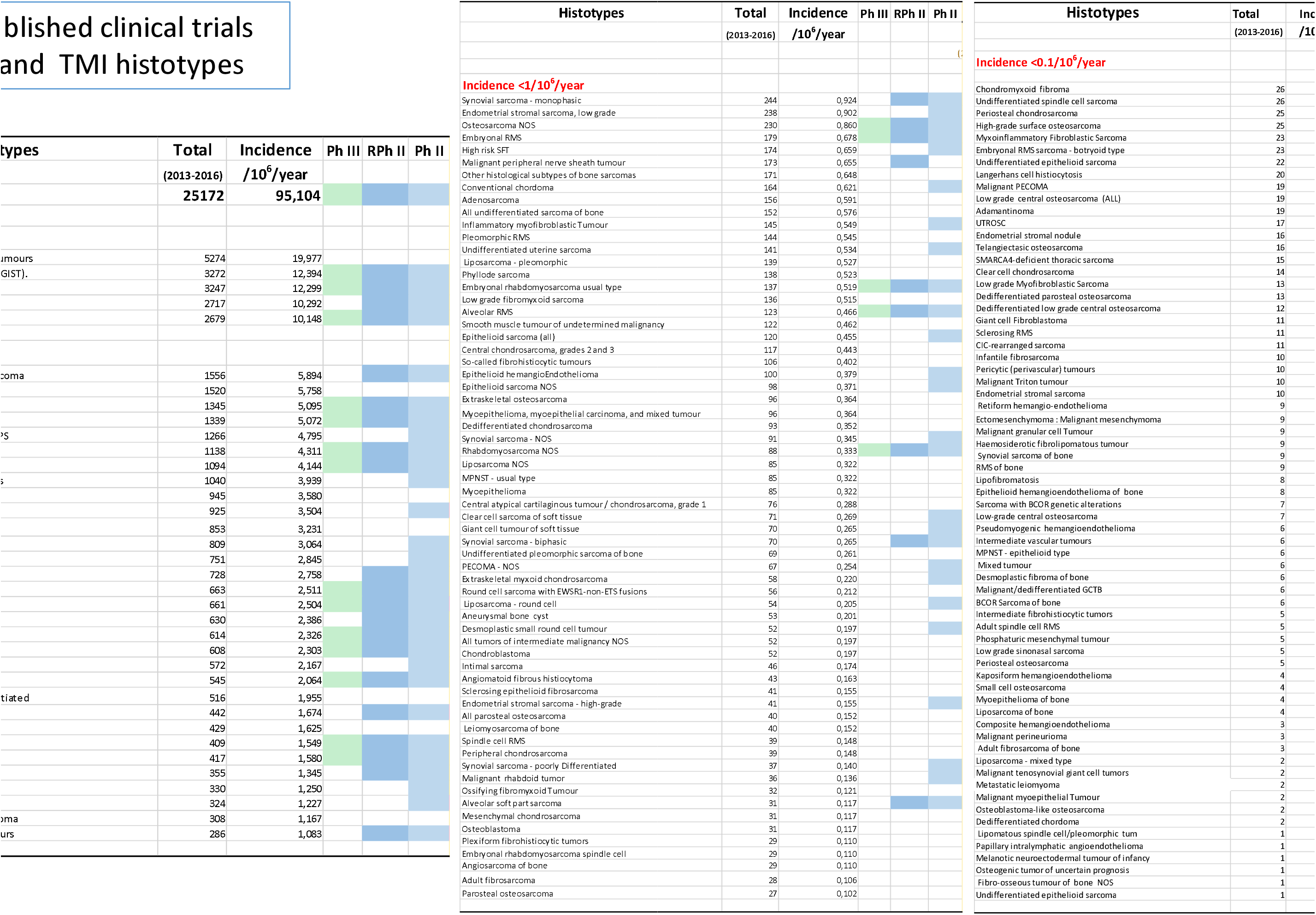
Published clinical trials in sarcoma and TMI histotypes. Tabular presentation of different sarcoma histotypes and groups of histotypes by decreasing order together with the documented published clinical trials in Pubmed: if phase III clinical trials are published, the box is highlighted in light green, if randomized phase II trials are published the box is highlighted in dark blue, if uncontrolled phase II trials are published the box is highlighted in light blue.

There were respectively 35, 63 and 66 different histological subtypes or groups (e.g. MPNST, or vascular sarcomas…) of sarcomas or TIM with an incidence ranging from 10 to 1/10^6^/year, 1-0.1/10^6^ per year, or <0.1/10^6^/year respectively.

These 3 groups gathered respectively 18542 (74%), 4766 (19%) and 568 (2%) of patients. The total number of patients in the different incidence groups exceeds the total number of patients of the series since some histological subtypes are listed as a group: for instance all liposarcoma as well as individual subtypes of liposarcomas (eg myxoid liposarcoma) are both listed.

A simple description of mean age, sex ratio, and site of the tumori s presented in table 4. It also shows the large clinical heterogeneity of these tumors with a mean age ranging from 5 years (infantile fibrosarcoma) to 78 (atypical fibroxanthoma), and a sex ratio from 0 (for sexual organs) to 153 for adenosarcoma.

**Table 4:** Age, gender, sites of histotypes

| Histotypes | Mean<br>Age | F/H<br>Ratio | Sites (%)* |  |  |  |  |  |  |  |
| --- | --- | --- | --- | --- | --- | --- | --- | --- | --- | --- |
|  |  |  | GI | Gyn | H&N | I. trnk | L. limb | Trnk w | U. limb | Others |
| Adamantinoma | 29,1 | 1,71 | 0,0 | 0,0 | 0,0 | 0,0 | 100,0 | 0,0 | 0,0 | 0,0 |
| Adenosarcoma | 61,3 | 51,00 | 1,9 | 89,7 | 0,0 | 3,8 | 0,6 | 0,6 | 0,0 | 3,2 |
| Adult fibrosarcoma | 69,5 | 1,15 | 0,0 | 7,1 | 7,1 | 10,7 | 17,9 | 28,6 | 17,9 | 10,7 |
| Adult spindle cell rhabdomyosarcoma | 52,4 | 0,25 | 0,0 | 0,0 | 20,0 | 0,0 | 20,0 | 40,0 | 20,0 | 0,0 |
| Alveolar rhabdomyosarcoma | 22,7 | 1,05 | 0,0 | 0,8 | 40,7 | 16,3 | 13,8 | 9,8 | 13,8 | 4,9 |
| Alveolar soft part sarcoma | 30,5 | 1,21 | 0,0 | 0,0 | 9,7 | 6,5 | 51,6 | 22,6 | 6,5 | 3,2 |
| Aneurysmal bone cyst | 30,2 | 1,12 | 0,0 | 0,0 | 3,8 | 1,9 | 39,6 | 32,1 | 20,8 | 1,9 |
| Angiomatoid fibrous histiocyoma | 27,1 | 0,87 | 0,0 | 0,0 | 7,0 | 11,6 | 37,2 | 18,6 | 25,6 | 0,0 |
| Angiosarcoma | 67,2 | 1,57 | 1,2 | 0,5 | 14,1 | 7,0 | 9,9 | 21,4 | 3,3 | 42,4 |
| Atypical cartilaginous Tumour/ChondroS G1 | 45 | 1,38 | 0,0 | 0,0 | 3,9 | 0,0 | 36,8 | 14,5 | 43,4 | 1,3 |
| Atypical fibroxanthoma | 78,7 | 0,20 | 0,2 | 0,0 | 12,8 | 0,0 | 1,6 | 82,1 | 1,6 | 1,6 |
| Atypical lipomatous tumor/WDLPS | 64,2 | 0,84 | 0,9 | 0,1 | 2,7 | 29,8 | 44,3 | 14,9 | 5,8 | 1,4 |
| BCOR sarcoma | 18,9 | 0,40 | 0,0 | 0,0 | 0,0 | 28,6 | 28,6 | 42,9 | 0,0 | 0,0 |
| Central chondrosarcoma | 56,4 | 0,98 | 0,0 | 0,0 | 7,7 | 13,7 | 35,0 | 17,1 | 26,5 | 0,0 |
| Chondroblastoma | 24,2 | 0,44 | 0,0 | 0,0 | 5,8 | 0,0 | 63,5 | 17,3 | 13,5 | 0,0 |
| Chondromyxoid fibroma | 34,1 | 0,73 | 0,0 | 0,0 | 3,8 | 0,0 | 42,3 | 30,8 | 23,1 | 0,0 |
| Chondrosarcoma NOS | 54,1 | 0,91 | 0,3 | 0,3 | 13,5 | 20,6 | 25,5 | 21,3 | 17,0 | 1,4 |
| Chordoma | 61,7 | 0,71 | 0,0 | 0,0 | 13,4 | 0,6 | 0,0 | 85,4 | 0,0 | 0,6 |
| CIC-DUX sarcoma | 24,1 | 0,38 | 9,1 | 0,0 | 9,1 | 0,0 | 45,5 | 27,3 | 0,0 | 9,1 |
| Clear cell chondrosarcoma | 42,5 | 0,27 | 0,0 | 0,0 | 0,0 | 7,1 | 64,3 | 7,1 | 21,4 | 0,0 |
| Clear cell sarcoma | 42,3 | 0,87 | 12,7 | 0,0 | 5,6 | 2,8 | 46,5 | 14,1 | 14,1 | 4,2 |
| Composite hemangioendothelioma | 33,3 | 0,50 | 0,0 | 0,0 | 0,0 | 0,0 | 33,3 | 66,7 | 0,0 | 0,0 |
| Conventional osteosarcoma | 32,6 | 0,80 | 0,0 | 0,0 | 12,5 | 3,1 | 60,2 | 14,4 | 9,6 | 0,2 |
| Dedifferentiated chondrosarcoma | 64 | 0,94 | 0,0 | 0,0 | 1,1 | 11,8 | 48,4 | 30,1 | 7,5 | 1,1 |
| Dedifferentiated chordoma | 53 | NA | 0,0 | 0,0 | 0,0 | 0,0 | 0,0 | 100,0 | 0,0 | 0,0 |
| Dedifferentiated low-grade central osteo | 36,2 | 1,40 | 0,0 | 0,0 | 0,0 | 0,0 | 83,3 | 8,3 | 8,3 | 0,0 |
| Dedifferentiated parosteal osteosarcoma | 43,8 | 2,25 | 0,0 | 0,0 | 7,7 | 0,0 | 76,9 | 7,7 | 7,7 | 0,0 |
| Dermatofibrosarcoma protuberans | 45 | 1,02 | 0,0 | 0,1 | 3,0 | 0,5 | 19,7 | 61,3 | 13,8 | 1,5 |
| Desmoid-type fibromatosis | 43,8 | 2,17 | 15,9 | 0,1 | 3,3 | 7,7 | 7,4 | 59,0 | 6,3 | 0,4 |
| Desmoplastic fibroma of bone | 27,5 | 1,00 | 0,0 | 0,0 | 0,0 | 0,0 | 66,7 | 16,7 | 16,7 | 0,0 |
| Desmoplastic round cell tumour | 24,3 | 0,30 | 3,8 | 0,0 | 0,0 | 84,6 | 0,0 | 0,0 | 0,0 | 11,5 |
| Embryonal rhabdomyosarcoma - botryoid type | 10,7 | 2,29 | 0,0 | 43,5 | 21,7 | 0,0 | 0,0 | 0,0 | 0,0 | 34,8 |
| Embryonal rhabdomyosarcoma - NOS | 20,3 | 0,71 | 0,0 | 16,7 | 41,7 | 8,3 | 0,0 | 8,3 | 0,0 | 25,0 |
| Embryonal rhabdomyosarcoma - spindle cell | 19 | 0,45 | 0,0 | 6,9 | 31,0 | 37,9 | 0,0 | 6,9 | 3,4 | 13,8 |
| Embryonal rhabdomyosarcoma - usual type | 14,4 | 0,57 | 0,9 | 4,4 | 35,4 | 38,9 | 3,5 | 3,5 | 0,9 | 12,4 |
| Endometrial stromal nodule | 50,7 | NA | 0,0 | 81,3 | 0,0 | 0,0 | 0,0 | 18,8 | 0,0 | 0,0 |
| Endometrial stromal sarcoma NOS | 56,2 | NA | 0,0 | 80,0 | 0,0 | 20,0 | 0,0 | 0,0 | 0,0 | 0,0 |
| Endometrial stromal sarcoma - high-grade | 60 | NA | 0,0 | 95,1 | 0,0 | 4,9 | 0,0 | 0,0 | 0,0 | 0,0 |
| Endometrial stromal sarcoma - low-grade | 53 | 211,00 | 0,5 | 88,7 | 0,0 | 9,4 | 0,0 | 0,5 | 0,0 | 0,9 |
| Epithelioid hemangioendothelioma | 52,1 | 1,56 | 2,0 | 0,0 | 9,0 | 11,0 | 14,0 | 14,0 | 5,0 | 45,0 |
| Epithelioid sarcoma | 40,1 | 0,85 | 1,0 | 6,1 | 2,0 | 12,2 | 22,4 | 19,4 | 29,6 | 7,1 |
| Ewing sarcoma | 26 | 0,68 | 1,0 | 0,5 | 5,7 | 16,6 | 28,2 | 33,6 | 7,8 | 6,7 |
| Extraskeletal myxoid chondrosarcoma | 58,2 | 0,81 | 0,0 | 0,0 | 0,0 | 1,7 | 58,6 | 27,6 | 8,6 | 3,4 |
| Extraskeletal osteosarcoma | 63,1 | 0,78 | 0,0 | 1,0 | 1,0 | 1,0 | 44,8 | 24,0 | 19,8 | 8,3 |
| Fibro-osseous tumour of bone NOS | 36 | NA | 0,0 | 0,0 | 0,0 | 0,0 | 0,0 | 100,0 | 0,0 | 0,0 |
| Fibrosarcomatous dermatofibrosarcoma prot. | 45,6 | 0,65 | 0,0 | 0,0 | 3,4 | 1,7 | 18,8 | 65,0 | 10,3 | 0,9 |
| Gastrointestinal stromal tumour (GIST), | 65,4 | 0,94 | 94,8 | 0,1 | 0,0 | 4,8 | 0,0 | 0,2 | 0,0 | 0,1 |
| Giant cell fibroblastoma | 22 | 0,38 | 0,0 | 0,0 | 0,0 | 0,0 | 54,5 | 45,5 | 0,0 | 0,0 |
| Giant cell tumour of bone | 37,8 | 1,13 | 0,0 | 0,0 | 0,9 | 0,0 | 58,3 | 14,4 | 25,1 | 1,3 |
| Giant cell tumour of soft tissues | 47,5 | 1,41 | 0,0 | 0,0 | 7,1 | 1,4 | 47,1 | 8,6 | 35,7 | 0,0 |
| Hemosiderotic fibrolipomatous tumour | 45,4 | 3,50 | 0,0 | 0,0 | 0,0 | 0,0 | 100,0 | 0,0 | 0,0 | 0,0 |
| High risk solitary fibrous tumour | 64,4 | 0,85 | 2,9 | 0,6 | 8,0 | 31,0 | 6,3 | 13,8 | 1,7 | 35,6 |
| High-grade surface osteosarcoma | 44,6 | 0,92 | 0,0 | 0,0 | 24,0 | 12,0 | 44,0 | 20,0 | 0,0 | 0,0 |
| Infantile fibrosarcoma | 5,9 | 2,33 | 10,0 | 0,0 | 20,0 | 10,0 | 20,0 | 20,0 | 0,0 | 20,0 |
| Inflammatory myofibroblastic tumour | 39,3 | 1,10 | 11,0 | 4,1 | 11,7 | 54,5 | 6,2 | 6,9 | 5,5 | 0,0 |
| Intermediate fibrohistiocytic tumours | 41 | 0,25 | 0,0 | 0,0 | 0,0 | 0,0 | 60,0 | 0,0 | 40,0 | 0,0 |
| Intermediate vascular tumours | 64,7 | 5,00 | 0,0 | 0,0 | 16,7 | 0,0 | 0,0 | 83,3 | 0,0 | 0,0 |
| Intimal sarcoma | 58,9 | 0,92 | 0,0 | 0,0 | 0,0 | 39,1 | 2,2 | 0,0 | 0,0 | 58,7 |
| Kaposi sarcoma | 65,8 | 0,22 | 1,1 | 0,0 | 3,2 | 1,1 | 65,8 | 11,5 | 13,4 | 4,1 |
| Kaposiform hemangioendothelioma | 6 | 3,00 | 0,0 | 0,0 | 0,0 | 0,0 | 50,0 | 50,0 | 0,0 | 0,0 |
| Langerhans cell histiocytosis | 29,5 | 4,00 | 0,0 | 0,0 | 10,0 | 5,0 | 5,0 | 75,0 | 5,0 | 0,0 |
| Leiomyosarcoma | 63,5 | 2,18 | 4,8 | 35,3 | 7,1 | 18,7 | 15,1 | 8,0 | 4,9 | 5,9 |
| Leiomyosarcoma - differentiated | 63,1 | 1,23 | 4,9 | 18,2 | 6,1 | 24,2 | 18,4 | 11,5 | 9,7 | 6,9 |
| Leiomyosarcoma - poorly-differentiated | 70,3 | 0,73 | 3,7 | 8,9 | 29,7 | 9,7 | 19,4 | 15,3 | 7,6 | 5,8 |
| Lipofibromatosis | 10,3 | 1,00 | 12,5 | 0,0 | 12,5 | 0,0 | 12,5 | 50,0 | 12,5 | 0,0 |
| Lipomatous spindle cell/pleomorphic tum | 33 | NA | 0,0 | 0,0 | 0,0 | 0,0 | 100,0 | 0,0 | 0,0 | 0,0 |
| Liposarcoma - dedifferentiated | 67,9 | 0,60 | 2,0 | 0,3 | 1,6 | 69,5 | 11,4 | 9,5 | 2,2 | 3,5 |
| Liposarcoma - mixed type | 61 | 1,00 | 0,0 | 0,0 | 0,0 | 50,0 | 50,0 | 0,0 | 0,0 | 0,0 |
| Liposarcoma - myxoid | 47,8 | 0,81 | 0,3 | 0,3 | 0,0 | 4,8 | 77,5 | 14,4 | 2,3 | 0,6 |
| Liposarcoma - NOS | 64,2 | 0,57 | 1,2 | 1,2 | 2,4 | 38,8 | 31,8 | 11,8 | 8,2 | 4,7 |
| Liposarcoma - pleomorphic | 63,1 | 0,78 | 0,7 | 0,7 | 3,6 | 15,1 | 38,1 | 22,3 | 15,8 | 3,6 |
| Liposarcoma - round cell | 49,2 | 0,59 | 0,0 | 0,0 | 0,0 | 14,8 | 63,0 | 14,8 | 7,4 | 0,0 |
| Low grade fibromyxoid sarcoma | 42,5 | 0,97 | 0,0 | 0,0 | 9,6 | 5,9 | 34,6 | 36,8 | 11,0 | 2,2 |
| Low grade myofibroblastic sarcoma | 40,5 | 1,17 | 7,7 | 0,0 | 46,2 | 0,0 | 23,1 | 23,1 | 0,0 | 0,0 |
| Low grade sinonasal sarcoma | 37,6 | 1,50 | 0,0 | 20,0 | 80,0 | 0,0 | 0,0 | 0,0 | 0,0 | 0,0 |
| Low-grade central osteosarcoma | 33,6 | 2,50 | 0,0 | 0,0 | 0,0 | 14,3 | 71,4 | 0,0 | 14,3 | 0,0 |
| Malignant glomus tumour | 54,2 | 0,67 | 20,0 | 10,0 | 10,0 | 10,0 | 30,0 | 0,0 | 20,0 | 0,0 |
| Malignant granular cell tumour | 46,1 | 1,25 | 0,0 | 0,0 | 0,0 | 0,0 | 0,0 | 55,6 | 44,4 | 0,0 |
| Malignant mesenchymoma | 61,1 | 1,25 | 0,0 | 11,1 | 0,0 | 11,1 | 33,3 | 0,0 | 33,3 | 11,1 |
| Malignant mixed tumor | 67,5 | NA | 25,0 | 75,0 | 0,0 | 0,0 | 0,0 | 0,0 | 0,0 | 0,0 |
| Malignant myoepithelial tumour | 49,5 | 1,00 | 0,0 | 0,0 | 0,0 | 0,0 | 0,0 | 50,0 | 50,0 | 0,0 |
| Malignant PECOMA | 60,1 | 1,71 | 15,8 | 21,1 | 0,0 | 26,3 | 10,5 | 10,5 | 0,0 | 15,8 |
| Malignant perineurioma | 46,3 | 2,00 | 0,0 | 0,0 | 0,0 | 0,0 | 33,3 | 33,3 | 0,0 | 33,3 |
| Malignant peripheral nerve sheath tumour | 46,4 | 0,86 | 0,6 | 0,0 | 11,6 | 17,3 | 24,9 | 31,2 | 9,8 | 4,6 |
| Malignant rhabdoid tumour | 24,3 | 0,89 | 2,8 | 8,3 | 13,9 | 16,7 | 5,6 | 16,7 | 0,0 | 36,1 |
| Malignant tenosynovial giant cell tumour | 68,5 | 0,00 | 0,0 | 0,0 | 0,0 | 50,0 | 0,0 | 50,0 | 0,0 | 0,0 |
| Malignant Triton tumour | 34,7 | 1,00 | 0,0 | 0,0 | 30,0 | 40,0 | 0,0 | 30,0 | 0,0 | 0,0 |
| Malignant/dedifferentiated giant cell tumor of the bone | 40 | 1,00 | 0,0 | 0,0 | 0,0 | 0,0 | 80,0 | 0,0 | 20,0 | 0,0 |
| Melanotic neuroectodermal tumour of infa | 38 | NA | 0,0 | 0,0 | 0,0 | 100,0 | 0,0 | 0,0 | 0,0 | 0,0 |
| Mesenchymal chondrosarcoma | 34,9 | 0,72 | 0,0 | 0,0 | 29,0 | 12,9 | 29,0 | 25,8 | 0,0 | 3,2 |
| Metastatic leiomyoma | 39 | NA | 0,0 | 0,0 | 0,0 | 50,0 | 0,0 | 50,0 | 0,0 | 0,0 |
| Mixed tumour | 66 | NA | 0,0 | 0,0 | 0,0 | 50,0 | 50,0 | 0,0 | 0,0 | 0,0 |
| MPNST - epithelioid type | 43,2 | 2,00 | 0,0 | 0,0 | 0,0 | 16,7 | 16,7 | 50,0 | 16,7 | 0,0 |
| MPNST - usual type | 45,4 | 0,77 | 1,2 | 1,2 | 14,1 | 12,9 | 27,1 | 27,1 | 11,8 | 4,7 |
| Myoepithelioma | 50,6 | 0,89 | 0,0 | 0,0 | 3,5 | 3,5 | 37,6 | 27,1 | 25,9 | 2,4 |
| Myxofibrosarcoma | 68,9 | 0,70 | 0,2 | 0,0 | 2,7 | 2,1 | 46,0 | 18,6 | 28,4 | 2,1 |
| Myxoinflammatory fibroblastic sarcoma | 54,3 | 0,53 | 0,0 | 0,0 | 0,0 | 0,0 | 39,1 | 0,0 | 60,9 | 0,0 |
| Osseous tumour rich in giant cell NOS | 40,5 | 1,00 | 0,0 | 0,0 | 0,0 | 0,0 | 50,0 | 50,0 | 0,0 | 0,0 |
| Ossifying fibromyxoid tumour | 49,8 | 1,13 | 0,0 | 0,0 | 9,4 | 6,3 | 12,5 | 37,5 | 34,4 | 0,0 |
| Osteoblastoma | 26,5 | 0,48 | 0,0 | 0,0 | 3,2 | 0,0 | 19,4 | 58,1 | 19,4 | 0,0 |
| Osteoblastoma-like osteosarcoma | 29 | NA | 0,0 | 0,0 | 0,0 | 0,0 | 100,0 | 0,0 | 0,0 | 0,0 |
| Osteogenic tumor of uncertain prognosis | 22 | NA | 0,0 | 0,0 | 0,0 | 0,0 | 0,0 | 100,0 | 0,0 | 0,0 |
| Osteosarcoma NOS | 38,3 | 0,64 | 0,0 | 0,0 | 15,7 | 2,6 | 51,3 | 19,1 | 9,1 | 2,2 |
| Papillary intralymphatic angioendothelio | 13 | NA | 0,0 | 0,0 | 0,0 | 0,0 | 0,0 | 100,0 | 0,0 | 0,0 |
| Parosteal osteosarcoma | 33,6 | 2,86 | 0,0 | 0,0 | 0,0 | 3,7 | 85,2 | 0,0 | 11,1 | 0,0 |
| PECOMA - NOS | 55,7 | 3,79 | 11,9 | 25,4 | 3,0 | 41,8 | 6,0 | 4,5 | 1,5 | 6,0 |
| Periosteal chondrosarcoma | 41,3 | 1,08 | 4,0 | 0,0 | 0,0 | 8,0 | 32,0 | 24,0 | 32,0 | 0,0 |
| Periosteal osteosarcoma | 19,8 | 4,00 | 0,0 | 0,0 | 0,0 | 20,0 | 80,0 | 0,0 | 0,0 | 0,0 |
| Peripheral chondrosarcoma | 37,3 | 0,63 | 0,0 | 0,0 | 0,0 | 10,3 | 33,3 | 33,3 | 23,1 | 0,0 |
| Phosphaturic mesenchymal tumour | 55,8 | 0,67 | 0,0 | 0,0 | 0,0 | 0,0 | 40,0 | 60,0 | 0,0 | 0,0 |
| Phyllodes sarcoma | 51,3 | 137,00 | 0,0 | 0,0 | 0,0 | 1,4 | 0,0 | 13,0 | 0,0 | 85,5 |
| Pleomorphic rhabdomyosarcoma | 67 | 0,58 | 1,4 | 7,6 | 5,6 | 11,8 | 36,1 | 19,4 | 11,1 | 6,9 |
| Plexiform fibrohistiocytic tumour | 23,1 | 1,23 | 0,0 | 0,0 | 6,9 | 3,4 | 34,5 | 24,1 | 31,0 | 0,0 |
| Pseudomyogenic hemangioendothelioma | 37,3 | 1,00 | 0,0 | 0,0 | 0,0 | 0,0 | 50,0 | 50,0 | 0,0 | 0,0 |
| Retiform hemangioendothelioma | 40,2 | 0,80 | 0,0 | 0,0 | 0,0 | 0,0 | 22,2 | 44,4 | 33,3 | 0,0 |
| Rhabdomyosarcoma - NOS | 41,7 | 0,80 | 1,1 | 13,6 | 17,0 | 23,9 | 11,4 | 4,5 | 5,7 | 22,7 |
| Sclerosing epithelioid fibrosarcoma | 55,8 | 1,05 | 0,0 | 0,0 | 9,8 | 17,1 | 14,6 | 41,5 | 9,8 | 7,3 |
| Sclerosing rhabdomyosarcoma | 45,2 | 0,38 | 0,0 | 0,0 | 9,1 | 0,0 | 72,7 | 0,0 | 9,1 | 9,1 |
| Small cell osteosarcoma | 21,8 | 0,33 | 0,0 | 0,0 | 0,0 | 25,0 | 25,0 | 0,0 | 25,0 | 25,0 |
| SMARCA4-deficient thoracic sarcoma | 48,3 | 0,25 | 13,3 | 0,0 | 0,0 | 40,0 | 0,0 | 0,0 | 0,0 | 46,7 |
| Smooth muscle tumour of undetermined mal | 50,6 | 4,30 | 6,6 | 59,8 | 0,8 | 15,6 | 5,7 | 7,4 | 4,1 | 0,0 |
| Solitary fibrous tumour | 58 | 1,25 | 1,2 | 1,1 | 15,4 | 21,0 | 13,3 | 18,6 | 4,3 | 25,0 |
| Spindle cell rhabdomyosarcoma | 38,8 | 0,63 | 5,1 | 2,6 | 17,9 | 20,5 | 28,2 | 10,3 | 12,8 | 2,6 |
| Suspicion of giant cell tumour of bone | 56,3 | 0,00 | 0,0 | 0,0 | 0,0 | 0,0 | 33,3 | 0,0 | 66,7 | 0,0 |
| Sarcoma NOS | 59,2 | 1,03 | 4,7 | 7,7 | 11,0 | 13,3 | 21,8 | 18,0 | 11,4 | 12,1 |
| Synovial sarcoma - biphasic | 41,2 | 0,84 | 1,4 | 0,0 | 5,7 | 5,7 | 52,9 | 17,1 | 10,0 | 7,1 |
| Synovial sarcoma - monophasic | 42,8 | 1,18 | 1,2 | 0,0 | 4,9 | 9,4 | 42,6 | 13,1 | 15,6 | 13,1 |
| Synovial sarcoma NOS | 45,6 | 1,22 | 0,0 | 1,1 | 5,5 | 8,8 | 34,1 | 13,2 | 16,5 | 20,9 |
| Synovial sarcoma - poorly differentiated | 45,2 | 0,85 | 0,0 | 0,0 | 5,4 | 10,8 | 29,7 | 18,9 | 8,1 | 27,0 |
| Telangiectasic osteosarcoma | 25,1 | 0,60 | 0,0 | 0,0 | 6,3 | 0,0 | 75,0 | 0,0 | 18,8 | 0,0 |
| Tumour of intermediate malignancy NOS | 47 | 1,14 | 5,8 | 3,8 | 13,5 | 13,5 | 21,2 | 19,2 | 21,2 | 1,9 |
| Undifferentiated epithelioid sarcoma | 70 | 0,47 | 0,0 | 0,0 | 13,6 | 18,2 | 22,7 | 13,6 | 27,3 | 4,5 |
| Undifferentiated pleomorphic sarcoma | 69,2 | 0,79 | 1,7 | 0,8 | 15,6 | 7,6 | 35,0 | 19,6 | 14,0 | 5,7 |
| Undifferentiated round cell sarcoma | 41,2 | 1,33 | 1,8 | 5,4 | 10,7 | 16,1 | 23,2 | 23,2 | 3,6 | 16,1 |
| Undifferentiated sarcoma | 66,7 | 0,89 | 1,4 | 3,3 | 15,8 | 13,0 | 29,5 | 21,4 | 8,8 | 6,7 |
| Undifferentiated sarcoma - NOS | 62,5 | 0,86 | 1,2 | 4,5 | 23,2 | 14,4 | 18,9 | 18,2 | 6,1 | 13,5 |
| Undifferentiated spindle cell sarcoma | 64,1 | 0,79 | 1,6 | 2,3 | 19,8 | 11,7 | 23,7 | 19,8 | 9,7 | 11,4 |
| Undifferentiated uterine sarcoma | 63,8 | NA | 0,0 | 96,1 | 0,0 | 2,9 | 0,0 | 1,0 | 0,0 | 0,0 |
| Uterine tumour resembling ovarian sex co | 48,1 | NA | 5,9 | 94,1 | 0,0 | 0,0 | 0,0 | 0,0 | 0,0 | 0,0 |

\* : Sites : GI : gastrointestinal ; Gyn : Gynaecological sites ; H&N : head and neck ; I. trnk : Internal trunk ; L. Limb : lower limb ; Trnk w : trunk wall ; U. limb : upper limb

### Variable incidence of sarcoma histotypes over the 2013-2016 period

We investigated then the variability of the yearly incidence of these different tumors in the database. The analysis of variance of the observed incidence indicated a significant interaction between time and histology (p<0.001). Supplementary figure presents the eight histological subtypes whose yearly incidence was found to vary significantly between 2013 and 2016. Adenosarcoma, central chondrosarcoma, solitary fibrous tumour, endometrial stromal sarcoma - high-grade increaseover the4 years, while intimal sarcoma, Kaposi sarcoma, Liposarcoma - round cell, myoepithelioma apprear to decrease. Most have an incidence <1/10^6^ year. While for Kaposi sarcoma, this maybe related to the evolving epidemiology of an associated condition (HIV infection), the significance of these variations remain unclear and will deserve to be explored in other registries with a central review.

### Incidence of individual histotypes and published clinical trials

Figure 1 presents graphically, and in ranking order (decreasing), the incidence of the different histotypes and groups of histotypes. These were matched with the presence of a published clinical trial on Pubmed focused specifically on this histological group (eg liposarcoma) or specific histotype (e.g. pleomorphic liposarcoma). Phase III studies, randomized phase II studies, and non randomized phase II studies are indicated in green, dark blue and light blue respectively. An histological subtype is considered covered by a trial only if the trial includes a specific arm (phase II) or a specific strata (phase III) in this given histotype.

As expected, phase III trials are available mostly in histotypes or groups of histotypes with an incidence >1/10^6^ per year (Figure 1).

14 of 35 (40%) histotypes with an incidence >1/10^6^ had a dedicated phase III study vs 6 of 129 (4.6%) histotypes for sarcomas with a incidence <1/10^6^ (p<10^−6^). 20100 (79,7%) patients of the database had a specific histotype for which no phase III trial had been reported.

21 of 35 (60%) histotypes with an incidence >1/10^6^ had a dedicated randomized phase II study vs 10 of 129 (7.7%) histotypes for sarcomas with a incidence <1/10^6^ (p<10^−10^)). 13154 (52.1%) patients of the database had a specific histotype for which no randomized phase II trial had been reported.

Twenty-eight of 35 (80%) histotypes with an incidence >1/10^6^ had a dedicated phase III study vs 36 of 129 (27.9%) histotypes for sarcomas with a incidence <1/10^6^ (p<10^−8^). 6516 (25.8%) patients of the database had a specific histotype for which no phase II trial had been reported.

## Discussion

The objective of this work was to describe the incidence of individual histological subtypes of sarcomas and TIM according to the most recent WHO classification. These cases were collected from the single NETSARC+ database, gathering the previous RREPS, RESOS and NETSARC databases (netsarc.org).

This work supported by the French NCI allowed therefore to measure the incidence of sarcomas and TIM in a nationwide database, close to exhaustivity given the stringent criteria of central pathology review in place since 2010. Since 2013, the number of patients included in the database per year is relatively stable suggesting that this is close to exhaustivity. We stopped the description on year 2016, since years 2017 to 2019 are still being monitored by the NETSARC+ now.

The first important observation is that the incidence of these tumors is larger than previously reported in each of these 4 years (1-15). Recently published data from countries in 4 continents reported an overall incidence ranging from 3 to 7.7/10^6^/year. The results of these studies are also heterogenous in terms of respective proportion of the groups of histotypes, ranging from 4 to 20% for undifferentiated sarcomas for instance. Taken together these observations suggest that mandatory central pathology review, results in a higher than reported incidence of almost all subtypes.

The present work also confirm that sarcoma histotype is a highly fragmented group of diseases, whose individual incidences may range from 10/10^6^ to less than 0.01/10^6^, ie a >1000-fold difference in incidence for tumors altogether considered as rare according to the international classification.

This is also a highly heterogenous group of tumors in terms of clinical presentations as shown by the diversity of sex ratio and mean age for diagnosis. Each of these entities should therefore benefit from a specific research programs to describe their natural history as well as the impact of current treatment on their disease course. This require a coordinated effort, worldwide, to achieve this goal given the rarity of certain histotypes. This is currently being conduction by intergroup studies, and international networks such as EURACAN. This work also confirms the important of national registries to investigate these rare subtypes.

An intriguing observation is that the incidence of these tumor may vary over time, and this was observed to be significant for 8 histotypes. While etiologic reasons may account fgor the reduction of Kaposi sarcoma in this time period, there is no obvious explanation for the seven other histotypes. Given the stability of the pathology team over this period this is not likely resulting from the pathology review. Epidemiological studies in other countries may be useful to confirm these variations, which may guide research on etiology of these most often rare sarcomas and TIM.

Another observation, expected by clinicians, is the link between the incidence and the availability of published prospective clinical research work to guide the management of individual subtypes. For decades the medical treatment of sarcomas used a one-size-fits-all approach for phase II to III clinical trials. Since 15 years, dedicated randomized phase II, III and phase II studies were implemented for specific histological subtypes, starting with GIST. This is more the exception of the rule though. The majority of histotypes described in this work, expecially those with an incidence under 1/10^6^/year have not had a dedicated phase II, randomized phase II or phase III clinical trial to guide clinical practice guidelines. This represents the majority of these patients for phase III, and still about 25% of the patients for phase II. This calls for a revision of the criterias to define standard treatment for such rare tumors where phase III are not feasible. Health authorities and reimbursement bodies should adapt their decisions on approval and reimbursement on the feasible level of evidence which could be reached for tumors with and incidence <1/10^e^6 per year in order not to discriminate against patients with rare cancers. It is important to remember that altogether patient with rare cancers represent 22% of all patient swith cnacers, and about 30% of the mortality due to cancer (26).

This study has many limitations. We can not exclude that patients may not reach our network despite the administrative incentive. This is true in particular for bone sarcoma and TIM (e.g. chondroblastomas, osteoblastoma, aneurysmatic bone cyst, etc..) which were collected more recently. The work must also adapt the the rapidly evolving classification of sarcomas, including now molecular subclassifications, which are not described here (for instance GIST, the novel NTRK sarcoma subgroup, BCOR, CIC-DUX4 sarcomas). To further explore the exhaustivity of the NETSARC+ bases, an ongoing project connects this base to the social security data base (SNDS) the single payer in France covering all citizens for all diseases (the Deepsarc project). This should enable a further refinement of these numbers.

Nevertheless, the observation that the incidence of sarcomas and TIM was higher than previously reported in this work during this time period provides a valuable information for this group of tumors.

In conclusion, this nationwide registry describes the incidence of sarcoma and TIM at a nationwide level over a 4 year period, with a central sarcoma pathologist expert review. It provides a benchmark for comparison with other registries worldwide and confirm the limitations of clinical research in sarcomas with an incidence <10^6^ per year. The observation of variable incidence for specific histological suibtype is intringuing and should be compared with data from other countries. Geographical research on the distribution of these cases over the national territory are ongoing.

## Data Availability

All data are presented in the manuscript

## Legends to the Figures

**Supplementary Figure: Variable incidence of sarcoma subtypes between 2013 to 2016**

Presentation of the yearly variation of the eight different histotypes with significantly variable incidence in the period of observation.

## References

1. Fletcher CDM, JA Bridge, PCW Hogendoorn, Mertens F. Pathology and genetics of tumours of soft tissue and bone. World Health Organization. IARC Press: Lyon 2013.

2. Zahm SH, Fraumeni JF Jr. The epidemiology of soft tissue sarcoma. Semin Oncol. 1997; 24:504–14.

3. Clark MA, Fisher C, Judson I, Thomas JM. Soft-tissue sarcomas in adults. N Engl J Med. 2005; 353:701–11.

4. Alvegård T, Sundby Hall K, Bauer H, Rydholm A. The Scandinavian Sarcoma Group: 30 years’ experience. Acta Orthop Suppl. 2009; 80:1-104.

5. Mastrangelo G, Fadda E, Cegolon L, et al. A European project on incidence, treatment, and outcome of sarcoma. BMC Public Health. 2010;10:188.

6. Ducimetière F, Lurkin A, Ranchère-Vince D, et al. Incidence of sarcoma histotypes and molecular subtypes in a prospective epidemiological study with central pathology review and molecular testing. PLoS One. 2011;6:e20294.

7. Ray-Coquard I, Montesco MC, Coindre JM, Dei Tos AP, Lurkin A, Ranchère-Vince D, Vecchiato A, Decouvelaere AV, Mathoulin-Pélissier S, Albert S, Cousin P, Cellier D, Toffolatti L, Rossi CR, Blay JY. Sarcoma: concordance between initial diagnosis and centralized expert review in a population-based study within three European regions. Ann Oncol. 2012;23:2442–2449.

8. C.A. Stiller, A. Trama, D. Serraino, S. et al, The RARECARE Working Group Descriptive epidemiology of sarcomas in Europe: Report from the RARECARE project. European Journal of Cancer 2013;49:684–695.

9. Gatta G, Capocaccia R, Botta L, Mallone S, De Angelis R, Ardanaz E, Comber H, Dimitrova N, Leinonen MK, Siesling S, van der Zwan JM, Van Eycken L, Visser O, Žakelj MP, Anderson LA, Bella F, Kaire I, Otter R, Stiller CA, Trama A; RARECAREnet working group. Burden and centralised treatment in Europe of rare tumours: results of RARECAREnet-a population-based study. Lancet Oncol. 2017; 18:1022–1039.

10. Trovik C, Bauer HCF, Styring E, Sundby Hall K, Vult Von Steyern F, Eriksson S, Johansson I, Sampo M, Laitinen M, Kalén A, Jónsson H Jr, Jebsen N, Eriksson M, Tukiainen E, Wall N, Zaikova O, Sigurðsson H, Lehtinen T, Bjerkehagen B, Skorpil M, Egil Eide G, Johansson E, Alvegard TA. The Scandinavian Sarcoma Group Central Register: 6,000 patients after 25 years of monitoring of referral and treatment of extremity and trunk wall soft-tissue sarcoma. Acta Orthop. 2017; 88:341–347.

11. Martin-Broto J, Hindi N, Cruz J, Martinez-Trufero J, Valverde C, De Sande LM, Sala A, Bellido L, De Juan A, Rubió-Casadevall J, Diaz-Beveridge R, Cubedo R, Tendero O, Salinas D, Gracia I, Ramos R, Baguè S, Gutierrez A, Duran-Moreno J, Lopez-Pousa A. Relevance of Reference Centers in Sarcoma Care and Quality Item Evaluation: Results from the Prospective Registry of the Spanish Group for Research in Sarcoma (GEIS). Oncologist. 2019; 24:e338–e346.

12. Kollár A, Rothermundt C, Klenke F, Bode B, Baumhoer D, Arndt V, Feller A; NICER Working Group. Incidence, mortality, and survival trends of soft tissue and bone sarcoma in Switzerland between 1996 and 2015. Cancer Epidemiol. 2019; 63:101596.

13. Ressing M, Wardelmann E, Hohenberger P, Jakob J, Kasper B, Emrich K, Eberle A, Blettner M, Zeissig SR. Strengthening health data on a rare and heterogeneous disease: sarcoma incidence and histological subtypes in Germany. BMC Public Health. 2018 Feb 12;18(1):235. doi: 10.1186/s12889-018-5131-4. PubMed PMID: 29433465; PubMed Central PMCID: PMC5809940.

14. Yang Z, Zheng R, Zhang S, Zeng H, Li H, Chen W. Incidence, distribution of histological subtypes and primary sites of soft tissue sarcoma in China. Cancer Biol Med. 2019; 16:565–574.. Bessen T, Caughey GE, Shakib S, Potter JA, Reid J, Farshid G, Roder D, Neuhaus SJ. A population-based study of soft tissue sarcoma incidence and survival in Australia: An analysis of 26,970 cases. Cancer Epidemiol. 2019; 63:101590.

15. Gage MM, Nagarajan N, Ruck JM, Canner JK, Khan S, Giuliano K, Gani F, Wolfgang C, Johnston FM, Ahuja N. Sarcomas in the United States: Recent trends and a call for improved staging. Oncotarget. 2019; 10:2462–2474.

16. Lurkin A, Ducimetière F, Vince DR, Decouvelaere AV, Cellier D, Gilly FN, Salameire D, Biron P, de Laroche G, Blay JY, Ray-Coquard I. Epidemiological evaluation of concordance between initial diagnosis and central pathology review in a comprehensive and prospective series of sarcoma patients in the Rhone-Alpes region. BMC Cancer. 2010; 10:150.

17. Casali PG, Bielack S, Abecassis N, et al; ESMO Guidelines Committee, PaedCan and ERN EURACAN. Bone sarcomas: ESMO-PaedCan-EURACAN Clinical Practice Guidelines for diagnosis, treatment andfollow-up. Ann Oncol. 2018;29 (Supplement_4):iv79–iv95.

18. Casali PG, Abecassis N, Bauer S, et al. ESMO Guidelines Committee and EURACAN. Gastrointestinal stromal tumours: ESMO-EURACAN Clinical Practice Guidelines for diagnosis, treatment and follow-up. Ann Oncol. 2018;29(Supplement_4):iv68–iv78.

19. Casali PG, Abecassis N, Bauer S, et al; ESMO Guidelines Committee and EURACAN. Soft issue and visceral sarcomas: ESMO-EURACAN Clinical Practice Guidelines for diagnosis, treatment and follow-up. Ann Oncol. 2018;29(Supplement_4):iv51–iv67.

20. von Mehren M, Randall RL, Benjamin RS, et al. Soft Tissue Sarcoma, Version 2.2016, NCCN Clinical Practice Guidelines in Oncology. J Natl Compr Canc Netw. 2016;14:758–86.

21. Dangoor A, Seddon B, Gerrand C, Grimer R, Whelan J, Judson I. UK guidelines for the management of soft tissue sarcomas. Clin Sarcoma Res. 2016;6–20.

22. Perrier L, Rascle P, Morelle M, Toulmonde M, Ranchere Vince D, Le Cesne A, Terrier P, Neuville A, Meeus P, Farsi F, Ducimetière F, Blay JY, Ray Coquard I, Coindre JM. The cost-saving effect of centralized histological reviews with soft tissue and visceral sarcomas, GIST, and desmoid tumors: The experiences of the pathologists of the French Sarcoma Group. PLoS One. 2018 Apr 5;13(4):e0193330

23. Perrier L, Buja A, Mastrangelo G, Vecchiato A, Sandonà P, Ducimetière F, Blay JY, Gilly FN, Siani C, Biron P, Ranchère-Vince D, Decouvelaere AV, Thiesse P, Bergeron C, Dei Tos AP, Coindre JM, Rossi CR, Ray-Coquard I. Clinicians’ adherence versus non adherence to practice guidelines in the management of patients with sarcoma: a cost-effectiveness assessment in two European regions. BMC Health Serv Res. 2012; 12:82.

24. Blay JY, Soibinet P, Penel N, Bompas E, Duffaud F, Stoeckle E, Mir O, Adam J, Chevreau C, Bonvalot S, Rios M, Kerbrat P, Cupissol D, Anract P, Gouin F, Kurtz JE, Lebbe C, Isambert N, Bertucci F, Toumonde M, Thyss A, Piperno-Neumann S, Dubray-Longeras P, Meeus P, Ducimetière F, Giraud A, Coindre JM, Ray-Coquard I, Italiano A, Le Cesne A. Improved survival using specialized multidisciplinary board in sarcoma patients. Ann Oncol. 2017; 28:2852–2859.

25. Blay JY, Honoré C, Stoeckle E, Meeus P, Jafari M, Gouin F, Anract P, Ferron G, Rochwerger A, Ropars M, Carrere S, Marchal F, Sirveaux F, Di Marco A, Le Nail LR, Guiramand J, Vaz G, Machiavello JC, Marco O, Causeret S, Gimbergues P, Fiorenza F, Chaigneau L, Guillemin F, Guilloit JM, Dujardin F, Spano JP, Ruzic JC, Michot A, Soibinet P, Bompas E, Chevreau C, Duffaud F, Rios M, Perrin C, Firmin N, Bertucci F, Le Pechoux C, Le Loarer F, Collard O, Karanian-Philippe M, Brahmi M, Dufresne A, Dupré A, Ducimetière F, Giraud A, Pérol D, Toulmonde M, Ray-Coquard I, Italiano A, Le Cesne A, Penel N, Bonvalot S; NETSARC/REPPS/RESOS and French Sarcoma Group–Groupe d’Etude des Tumeurs Osseuses (GSF-GETO) Networks. Surgery in reference centers improves survival of sarcoma patients: a nationwide study. Ann Oncol. 2019; 30:1143–1153.

26. Gatta G, van der Zwan JM, Casali PG, Siesling S, Dei Tos AP, Kunkler I, Otter R, Licitra L, Mallone S, Tavilla A, Trama A, Capocaccia R; RARECARE working group. Rare cancers are not so rare: the rare cancer burden in Europe. Eur J Cancer. 2011; 47:2493–511.

